# Estimating Heritability of Glycaemic Response to Metformin using Nationwide Electronic Health Records and Population-Sized Pedigree

**DOI:** 10.1101/2020.08.24.20173922

**Authors:** Iris N. Kalka, Amir Gavrieli, Smadar Shilo, Hagai Rossman, Nitzan Shalom Artzi, Eran Segal

**Affiliations:** Department of Computer Science and Applied Mathematics, Weizmann Institute of Science, Rehovot, Israel; Department of Molecular Cell Biology, Weizmann Institute of Science, Rehovot, Israel; Pediatric Diabetes Unit, Ruth Rappaport Children’s Hospital, Rambam Healthcare Campus, Haifa, Israel

## Abstract

Variability of response to medication is a well known phenomenon, determined by both environmental and genetic factors. Understanding the heritable component of the response to medication is of great interest but challenging due to several reasons, including small study cohorts and computational limitations. Here, we studied the heritability of variation in the glycaemic response to metformin, first-line therapeutic agent for type 2 diabetes (T2D), by leveraging 17 years of electronic health records (EHR) data from Israel’s largest healthcare service provider, consisting of over five million patients of diverse ethnicities and socio-economic background. Our cohort consisted of 74,871 T2D patients treated with metformin, with an accumulated number of 1,358,776 HbA1C measurements and 323,260 metformin prescriptions. We estimated the explained variance of glycated hemoglobin (HbA1c%) reduction due to heritability by constructing a six-generation population-size pedigree from pedigree information linked to medical health records. Using a Linear Mixed Model-based framework, a common-practice method for heritability estimation, we calculated a heritability measure of *h*^2^ = 10.5% (95% CI, 3.5%-17.5%) for absolute reduction of HbA1c% after metformin treatment, which remained unchanged after adjusting for pre-treatment HbA1c%, and *h*^2^ = 12.2% (95% CI, 5.2%-19.3%) for proportional reduction in HbA1c%. To the best of our knowledge, our work is the first to estimate heritability of drug response using EHR data. We demonstrated that while response to metformin treatment has a heritable component, most of the variation is likely due to other factors, further motivating non-genetic analyses aimed at unraveling metformin’s mechanism of action.

## Introduction

During the past three decades there has been a two fold increase in the prevalence of diabetes in the general population (WHO), currently estimated to afflict one in every 16 adults ^1^. Type 2 diabetes (T2D), which accounts for approximately 90% of the total diabetic population, is a major cause of morbidity and is among the top 10 mortality causes in adults^2,3^.

Metformin is the first-line oral agent for lowering blood sugar levels in T2D patients. Through inhibition of hepatic glucose production it reduces intestinal glucose absorption, and improves both glucose uptake and its utilization ^4^. The significant role of metformin in diabetes management is particularly remarkable since its mechanism is still not fully understood ^5-7^.

Glycaemic response to metformin is varied across patients ^6-7^, and remains unexplained by individual features. Some variation can be accounted for by personal characteristics including sex, age and BMI, as well as features describing treatment strategies such as dosage and adherence ^8^. In addition, a small fraction of the response variability is attributed to genetic variants, providing motivation to further explore heritable variance in metformin response ^9^.

Medication response variations are widely agreed upon to be determined by the interplay of environmental and genetic factors ^11,12^. The effect of heritable factors has been suggested as early as 1908 ^10^. This notion led to the development of pharmacogenomics, which investigates genetic variants that account for differential drug responses and personal response to treatments ^13^.

Traditionally heritability estimates are deciphered through twins and family studies, however, those are difficult to construct in the context of medication response. Drug response data, same diagnosis and similar treatment are rarely available in multiple family members ^14,15^. Moreover, because close relatives often share environment and not only genetics, such studies have difficulties in separating the genetic and environmental effects.

Other types of studies estimating the effect of genetic variability in drug responses rely on small cohorts undergoing costly genetic tests and use genetic relatedness estimation methods ^16-19^. Due to the sizes of such cohorts, these estimates tend to have low statistical power and do not represent the true distribution of the population ^20^.

In studies bypassing genetic tests, such as family-linkage studies, information is highly sparse and determining the response to medication by genetic and environmental factors is computationally challenging. Epigenetics may also play a role in the response to medication making the task even harder ^21^.

Metformin’s effect is routinely measured through glycaemic control assessments using either fasting glucose or HbA1c% ^22^. The latter is an indicator of blood sugar levels over the course of three months^23^, making it more reliable than the former, which is a snapshot of a single time point. Moreover, fasting glucose is affected by the strictness of fasting prior to blood test, a measure which is not recorded, making fasting glucose more prone to mistakes.

In this study we used Electronic Health Records (EHRs) from Clalit Healthcare database, Israel’s largest healthcare service provider ^24^. This population-size EHR provides a real-world view of the internal variability in healthcare systems, where patients, diagnoses, and treatment plans vary considerably. In general, EHRs can contain medical information on millions of patients, however data is sparse and noisy and not cross-sectional ^25^. Combined with pedigree information this unique data allowed us to include family medical history of first order relatives and extended family members alike.

Today, heritability estimation is typically performed using Linear Mixed Models (LMMs) ^26^ and requires a kinship matrix, commonly computed from genetic information. Using the Sparse Cholesky Factorization (Sci-LMM) package^27^, a statistical modeling framework for analyzing population-size pedigrees, we constructed a relationship matrix and fitted the corresponding LMM solely from EHR information, without costly genetic testing. We estimated the heritability of absolute HbA1c% reduction in response to metformin to be 10.5% (95% CI 3.5%-17.5%) of the total explained variability.

## Methods

### Data

We used EHRs of Clalit Health Services (Clalit), Israel’s largest healthcare provider. Clalit’s data are heterogeneous in terms of geography and socioeconomics, including more than five million people (over half of Israel’s population) with longitudinal measurements dating back to 2002. EHRs are reflective of the members’ full clinical experience including diagnoses, lab tests results and medication prescribed and dispensed. Patients’ information is combined with pedigree information to provide demographics consisting of date of birth, sex, parental information and county of birth, from which ethnicity is inferred ^28^.

### Pedigree and kinship matrix construction

We obtained pedigree information through demographics of past and present patients as well as their parents, and then excluded cases where parental relationships and sex contradicted (e.g. a female father). We converted the entire pedigree to a directed graph using NetworkX ^29^, where nodes and edges corresponded to individuals and to parenthood respectively, and removed all edges of directed cycles, as these are not feasible ^30^.

Heritability estimates require a kinship matrix, also known as an Additive Relationship Matrix (ARM)^31^, measuring the proportion of identical genes between pairs of individuals. Using the Nadiv R package ^32^, we approximated the ARM solely from pedigree information, under the assumption that genes distribute uniformly, meaning each gene has an identical probability to be passed on ^33^. For every pair of individuals and a unique shortest path between them through a shared ancestor we increased their similarity by 2^−^*^l^* where *l* is the number of edges in the path (Figure S1a-c).

We decided against removing first-degree relatives in heritability estimates. Although some studies suggest it reduces estimation bias, we found it less relevant to our case ^34^ (Supplementary discussion 1).

### Identification of T2D patients

In Israel, T2D is diagnosed based on plasma glucose criteria, in accordance with The American Diabetes Association standard of care ^35^. Meeting any of the following criteria is sufficient for T2D diagnosis: (1) random plasma glucose ≥200 mg/dL (2) HbA1c% ≥6.5% (3) Two separate test samples of fasting plasma glucose ≥ 126 mg/dL following no caloric intake for at least eight hours (4) plasma glucose ≥ 200 mg/dL two hours after oral glucose tolerance test (OGTT).

Note that although fasting glucose could be used in diagnosis of T2D, data is inaccurate as some non-fasting patients take the test as well. Also, OGTT tests are not performed regularly in clinics, making us disregard the corresponding criterion.

In addition to identifying T2D patients through test results, we made use of diagnoses data. Including all patients diagnosed with T2D according to the appropriate International Classification of Diseases, Ninth Revision (ICD-9) codes ^36^ (Supplementary Table 2).

### Cohort definition

Our cohort constitutes T2D patients treated for diabetes with metformin only after diabetes diagnosis. We identified those from drug prescriptions with fifth level Anatomical Therapeutic Chemical (ATC) ^37^ code of ‘A10BA02’. We defined the first metformin prescription date for every patient as index-date^37^, yielding a single unique date per individual by which all other dates are measured.

To establish glycaemic response to metformin we used HbA1c% blood concentration before and after metformin treatment initiation (Figure S1d). We defined baseline (pretreatment) HbA1c% as the latest test occurring 90 days prior to 14 days post index-date. This interval was chosen as part of the balance between to ensure measurements are within a red blood cell life cycle and since metformin onset of action is within two weeks ^38^-^39^. To ensure stability of results, we estimate heritability on several baseline time intervals for the entire cohort (Supplementary Table 4). We define the on-treatment HbA1c% as the closest test to the index-date that is at least 90 days from both index-date and baseline HbA1c% date, indicating hemoglobin turning rate. We discarded on-treatment HbA1c% tests later than 180 days from index-date, as those are confounded by unmeasured variables. We defined the study participation period as the time from index-date or baseline measurement date, whichever preceded, until the on-treatment measurement date.

We ensured measuring the effect of metformin by further screening patients who were treated throughout the entire study participation period. We removed all patients who stopped metformin treatment before on-treatment HbA1c% test or who started taking metformin before being diagnosed with T2D, the majority of which were prescribed metformin while already diagnosed as pre-diabetic. We also exclude all patients who are prescribed any other anti-diabetic medication (ATC level 2 code of ‘A10’) apart from metformin to ensure the effect on HbA1c% levels can be attributed solely to metformin.

We further removed all patients with abnormal estimated Glomerular Filtration Rate (eGFR) who should not be treated according to medical guidelines ^39,40^. GFR is estimated using creatinine blood tests and reflects renal clearance and total clearance, which after oral administration of metformin decrease approximately in proportion to it ^41^ (Table 1).

**Table 1:** Inclusion exclusion criteria.

| Table 1: Inclusion exclusion criteria |  |
| --- | --- |
| Inclusion | Exclusion |
| Diabetic treated with metformin only after diagnosis. | Abnormal estimated Glomerular Filtration Rate (eGFR<30 mL/min/1.73m2) |
| Baseline HbA1c% test exists 90 days prior to 30 days after first metformin prescription | Treated for diabetes with non-metformin drugs (ATC2 is 'A10') |
| On-treatment HbA1c% test exists 90 to 180 days after first metformin prescription | Baseline HbA1c% and on-treatment HbA1c% tests <90 days apart |

### Glycaemic response outcomes

We defined three phenotypes commonly used in metformin pharmacogenetics studies for measuring the response to metformin; absolute, proportional and adjusted reduction in HbA1c%^16^. These were induced from the difference of the baseline and the on-treatment HbA1c% tests. Absolute reduction was defined as the absolute difference between on-treatment and baseline HbA1c%, proportional reduction was defined as the absolute reduction divided by the baseline HbA1c%, and adjusted reduction was defined as the residuals from the model’s outcome using the absolute reduction as the phenotype. Since Linear Mixed Models assume normal distribution we performed the Kolmogorov-Smirnov goodness of fit test for all three phenotypes ^43,44^.

### Height outcome

Being that heritability estimate of height is well established and agreed upon in the literature, we used it as a positive control to validate our methods and data. We gathered height measurements recorded at adulthood (age ≥ 18 years). For patients who had multiple measurements, we considered the latest measurement only. We removed outlier measurements where Z-score<4.

### Heritability estimation

We computed heritability with the Sci-LMM Python package, which constructs and works with large scale relationships matrices and fits them to the corresponding LMM within several hours. Our Identity By Descent (IBD) matrix (an identity-by-descent relationships based matrix) was the ARM computed from the entire pedigree^27^. We used Haseman-Elston regression to compute the heritability measure *h*^2^ and its 95% confidence interval^45^.

We constructed the following features used either as covariates for our regression model or as means of subsampling the cohort:

1. Demographics:

a. Year of birth
b. Age at index-date
c. Gender
d. BMI: note that since is considered heritable we did not use it as a covariate in our regression.
2. Measurements metadata:

a. Baseline to index gap: number of days between baseline-date to index-date
b. Index to on-treatment gap: number of days between index-date and on-treatment date
c. Baseline to on-treatment gap: number of months between on-treatment date and baseline-date.
d. Number of HbA1c% tests: the absolute number of HbA1c% tests performed up until on-treatment date
3. Lab test measurements:

a. Estimated Glomerular Filtration Rate (eGFR): eGFR was used MDRD GFR Equation^46^: *eGFR =* 186 × *creatinine* ^−1.154^ × *age*^−0.203^ where value is multiplied by 0.742 for females.
b. Baseline HbA1c%
4. Treatment metadata:

a. Average dosage: weighted average of metformin doses Σ*w_i_* × *p_i_/*Σ*p_i_* where *w_i_* is the number of pills per day prescribed in prescription *i*, and *p_i_* is the number of pills in prescription *i*. Only issued prescriptions were accounted for.
b. Adherence: since adherence is not reported, we capture it through four features representing the average number of days on metformin in four equal consecutive time intervals between index-date and on-treatment date. We assumed that all dispensed prescriptions were also consumed by patients.

## Results

### Cohort description

To estimate the heritability measure of response to metformin we extracted 721,123 T2D patients from Clalit Healthcare EHR database (Figure 1). Of these, we included only patients who had at least two HbA1c% test measurements, one before metformin treatment (baseline) and one after it (on-treatment). We excluded subjects treated with non-metformin diabetic drugs prior to on-treatment date, and those treated with metformin prior to diabetes diagnosis, maintaining a total of 74,871 patients.

**Figure 1:**
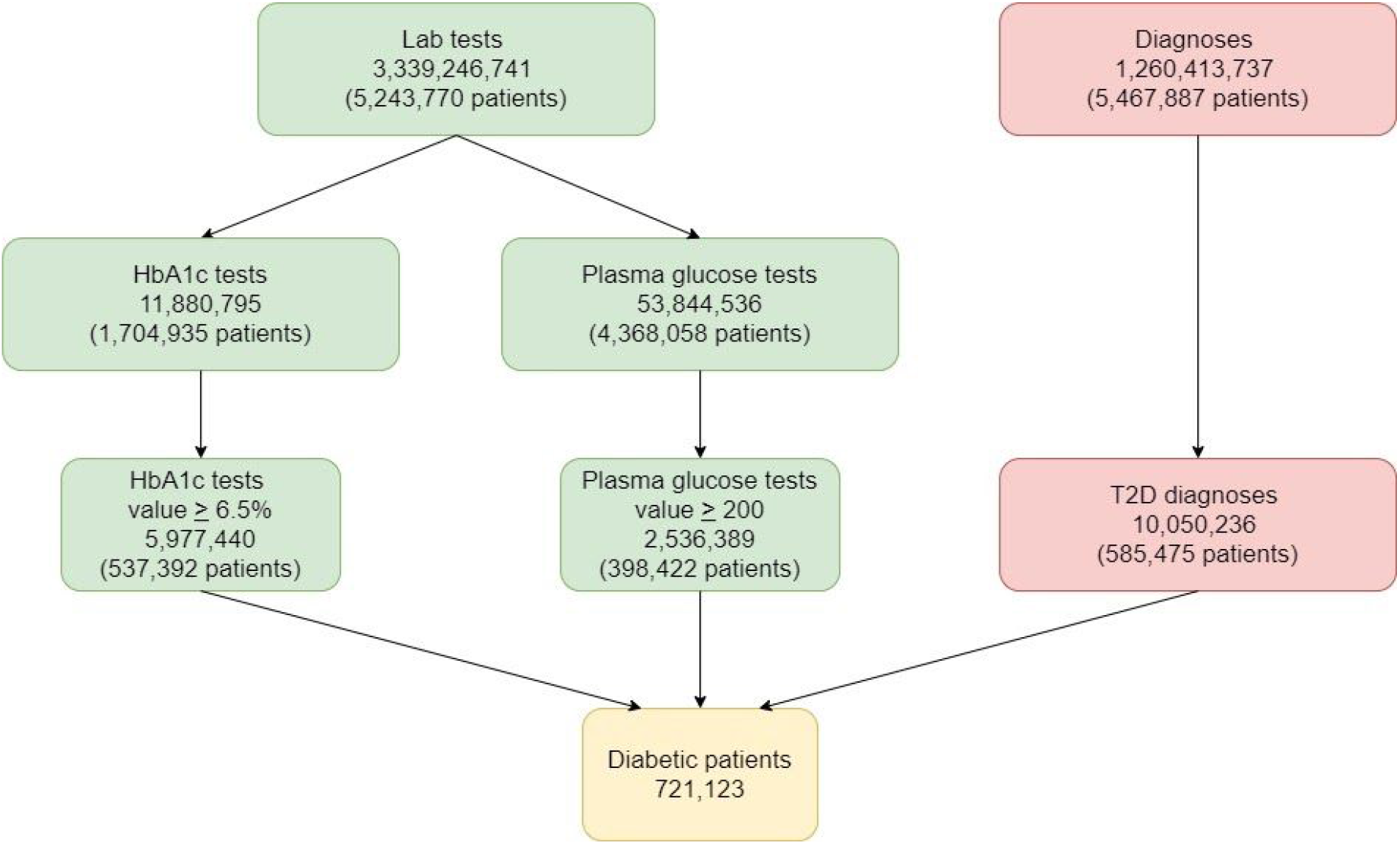
T2D patients selection. Patients were determined by fulfilling at least one of three criteria: (1) HbA1c% lab test value>6.5% (2) Plasma glucose lab test value>200mg/dL (3) T2D diagnosis. Green applies to lab test results, red to diagnoses and yellow to patients. In total 721,123 T2D patients were identified.

In total our cohort was balanced between genders, with 48.2% males. When comparing feature distribution between genders we found they all differ significantly, with the exception of average dose (e.g. average ages were 59.48 for males and 61.0 for females) (Table 2). We see therefore that although the two populations differ on every parameter, they receive similar treatments. Since they are all T2D patients, our cohort diverts from the general population by risk factors characteristics. In addition to being older and having an overweight BMI on average, patients had a baseline HbA1c% of 7.8% for males and 7.4% for females, nearly 1% over diabetic threshold HbA1c% value (6.5%). We identified that T2D patients had 1.85 first degree relatives on average also diagnosed with T2D. Out of the T2D patients, 32.9% had at least one first degree relative diagnosed with T2D.

**Table 2:**
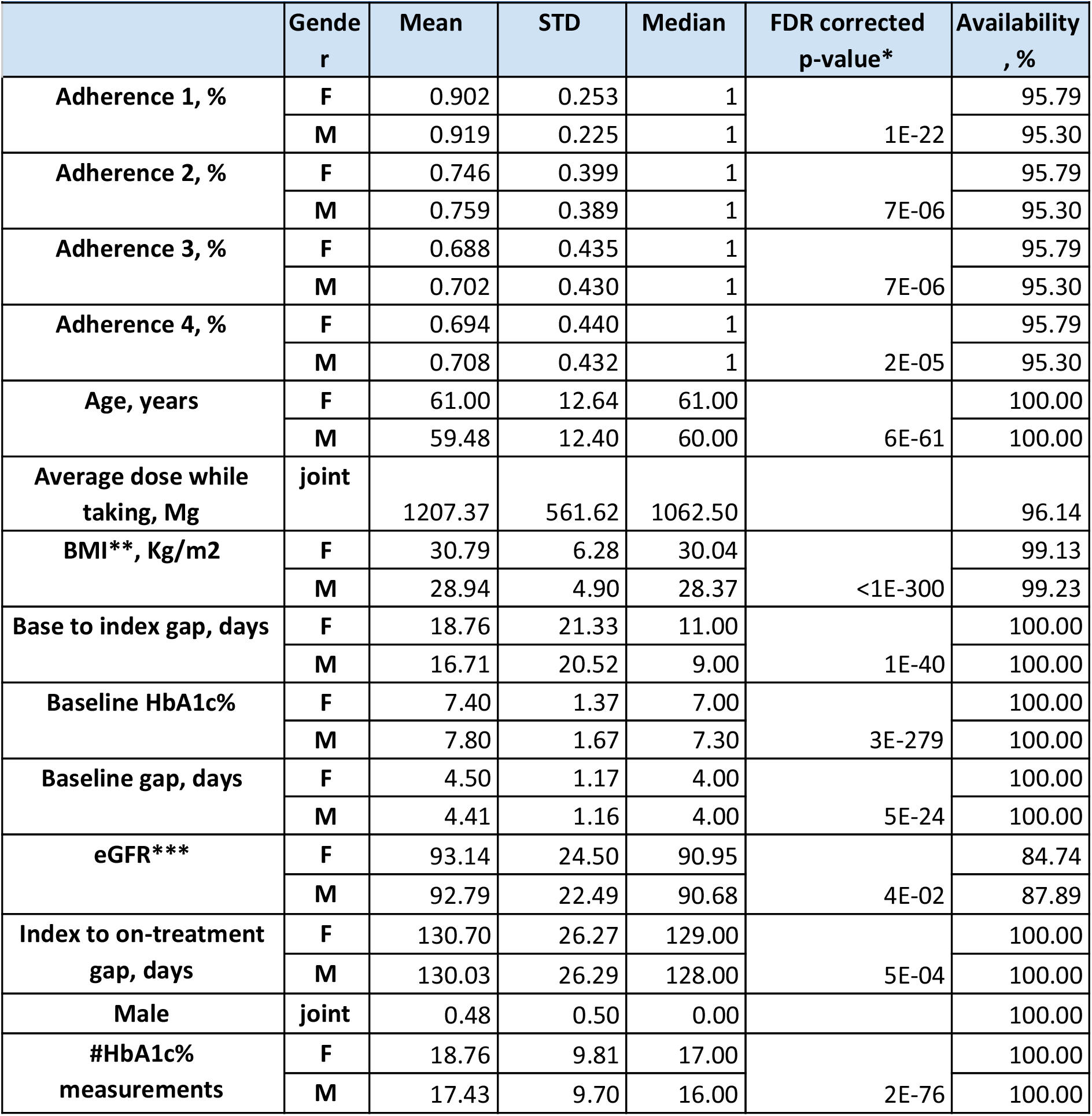
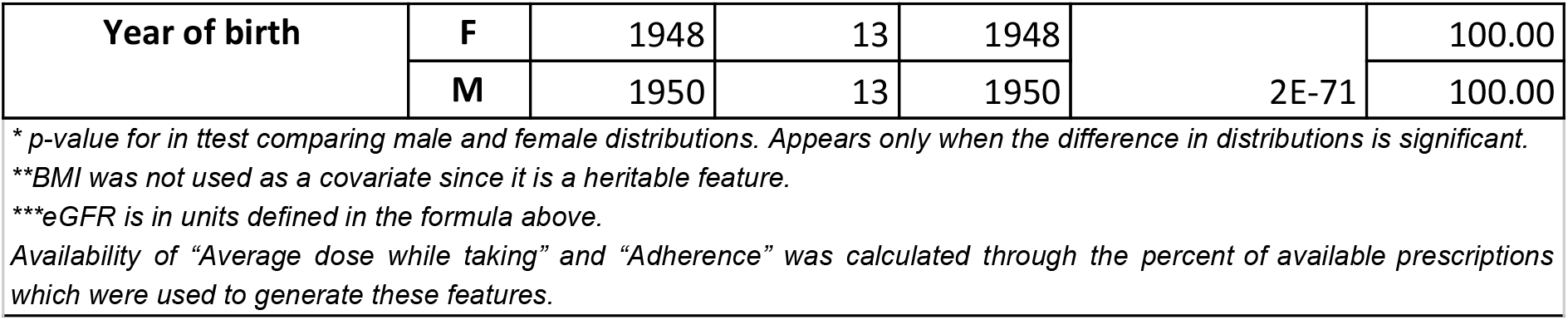
Baseline characteristics of the study cohort.

| Table 2: Baseline characteristics of the study cohort |  |  |  |  |  |  |
| --- | --- | --- | --- | --- | --- | --- |
|  | Gender | Mean | STD | Median | FDR corrected p-value* | Availability, % |
| Adherence 1, % | F | 0.902 | 0.253 | 1 | 1E-22 | 95.79 |
|  | M | 0.919 | 0.225 | 1 |  | 95.30 |
| Adherence 2, % | F | 0.746 | 0.399 | 1 | 7E-06 | 95.79 |
|  | M | 0.759 | 0.389 | 1 |  | 95.30 |
| Adherence 3, % | F | 0.688 | 0.435 | 1 | 7E-06 | 95.79 |
|  | M | 0.702 | 0.430 | 1 |  | 95.30 |
| Adherence 4, % | F | 0.694 | 0.440 | 1 | 2E-05 | 95.79 |
|  | M | 0.708 | 0.432 | 1 |  | 95.30 |
| Age, years | F | 61.00 | 12.64 | 61.00 | 6E-61 | 100.00 |
|  | M | 59.48 | 12.40 | 60.00 |  | 100.00 |
| Average dose while taking, Mg | joint | 1207.37 | 561.62 | 1062.50 |  | 96.14 |
| BMI**, Kg/m2 | F | 30.79 | 6.28 | 30.04 | <1E-300 | 99.13 |
|  | M | 28.94 | 4.90 | 28.37 |  | 99.23 |
| Base to index gap, days | F | 18.76 | 21.33 | 11.00 | 1E-40 | 100.00 |
|  | M | 16.71 | 20.52 | 9.00 |  | 100.00 |
| Baseline HbA1c% | F | 7.40 | 1.37 | 7.00 | 3E-279 | 100.00 |
|  | M | 7.80 | 1.67 | 7.30 |  | 100.00 |
| Baseline gap, days | F | 4.50 | 1.17 | 4.00 | 5E-24 | 100.00 |
|  | M | 4.41 | 1.16 | 4.00 |  | 100.00 |
| eGFR*** | F | 93.14 | 24.50 | 90.95 | 4E-02 | 84.74 |
|  | M | 92.79 | 22.49 | 90.68 |  | 87.89 |
| Index to on-treatment gap, days | F | 130.70 | 26.27 | 129.00 | 5E-04 | 100.00 |
|  | M | 130.03 | 26.29 | 128.00 |  | 100.00 |
| Male | joint | 0.48 | 0.50 | 0.00 |  | 100.00 |
| #HbA1c% measurements | F | 18.76 | 9.81 | 17.00 | 2E-76 | 100.00 |
|  | M | 17.43 | 9.70 | 16.00 |  | 100.00 |
| Year of birth | F | 1948 | 13 | 1948 | 2E-71 | 100.00 |
|  | M | 1950 | 13 | 1950 |  | 100.00 |

\* p-value for in ttest comparing male and female distributions. Appears only when the difference in distributions is significant.
\*\*BMI was not used as a covariate since it is a heritable feature.
\*\*\*eGFR is in units defined in the formula above.
Availability of "Average dose while taking" and "Adherence" was calculated through the percent of available prescriptions which were used to generate these features.

### Height heritability estimation

We first set out to validate our framework through estimation of height heritability, as it is a well-documented heritable measure ^48^. Such validation indicates whether the constructed pedigree could be used for heritability estimation, and also whether the Clalit’s EHR population is representative of the general population. Our estimation took into account two covariates, sex and year of birth, both of which are highly correlated with height regardless of heritable effects^49^.

We extracted height measurements of 11,670,689 adults from a total of 5,349,004 families with the largest family consisting of 4,310,015 adults. A Kolmogorov-Smirnov test indicated that height followed normal distribution (Supplementary Table 1) with a mean of 1.66 meters and a standard deviation of 0.10 meters (Table 3). We estimated the heritability measure of height to be *h*^2^ = 80.7% (95% CI 80.3%-81.1%), a value that is consistent with the literature^48^, thus validating both our approach and our dataset (Table 4).

**Table 3:** Cohort Phenotypes Statistics.

|  | Gender | Mean | Standard deviation | Median | FDR corrected p-value |
| --- | --- | --- | --- | --- | --- |
| HbA1c% absolute reduction | F | 0.68 | 1.23 | 0.40 | 6E-293 |
|  | M | 1.06 | 1.61 | 0.60 |  |
|  | joint | 0.86 | 1.44 | 0.50 |  |
| HbA1c% adjusted reduction | F | -0.12 | 0.89 | -0.12 | 1E-93 |
|  | M | 0.03 | 1.06 | -0.01 |  |
|  | joint | -0.05 | 0.98 | -0.07 |  |
| HbA1c% proportional reduction | F | 0.08 | 0.12 | 0.06 | 2E-285 |
|  | M | 0.11 | 0.15 | 0.08 |  |
|  | joint | 0.09 | 0.14 | 0.07 |  |
| Height, m | F | 1.60 | 0.07 | 1.60 | <1E-350 |
|  | M | 1.73 | 0.07 | 1.73 |  |
|  | joint | 1.66 | 0.10 | 1.66 |  |

\* Adjusted reduction is defined as the residuals from the model's outcome using the absolute reduction as the phenotype

**Table 4:** Heritability Estimates of Metformin Responses.

| Cut | HbA1c% absolute reduction |  | HbA1c% adjusted reduction |  | HbA1c% proportional reduction |  | Number of patients |
| --- | --- | --- | --- | --- | --- | --- | --- |
|  | h2 | CI | h2 | CI | h2 | CI |  |
| All cohort | 0.105 | [0.035,0.175] | 0.105 | [0.035,0.175] | 0.122 | [0.052,0.193] | 74,871 |
| Male | 0.159 | [0.012,0.305] |  |  | 0.173 | [0.025,0.321] | 36,098 |
| Female | 0.209 | [0.075,0.343] | 0.213 | [0.079,0.347] | 0.209 | [0.076,0.343] | 38,773 |

h2 estimates and their confidence intervals of our cohort including different subgroups. We only included subgroups which had positive h2 estimates with 95% confidence intervals.

### Metformin response characteristics

We computed three outcomes of response to newly metformin treated T2D patients from HbA1c% reduction: absolute, adjusted and proportional. We find that the mean absolute HbA1c% reduction is 0.86%, which concurs with known reductions after first time metformin treatment ^42^. It is important to note that response depends on treatment policy, as individuals with higher baseline HbA1c% receive higher doses of metformin which in turn result in larger HbA1c% reductions.

When observing the different phenotypes we find that they all differ significantly between males and females (Table 3). This led us to computing heritability estimates for each sex independently in addition to the entire cohort. Furthermore, we decided to compute estimates for additional subgroups within the population to better understand their independent heritable effect on metformin response.

### Heritability of response to metformin

We computed the heritability of HbA1c% reduction phenotypes on the entire cohort of 74,871 patients. For our calculations we used covariates of personal measures as well as treatment strategy measures (see Methods). We found that the heritability measure of absolute HbA1c% reduction is *h*^2^ = 10.5% (95% CI, 3.5%- 17.5%) (Table 4).

We examined whether our heritability estimates remain similar between different metformin response phenotypes. We found that the heritability estimate for the adjusted HbA1c% reduction is the same. This is expected, as the adjusted reduction values are the residuals from a model that is based on covariates. We find that the estimates for proportional HbA1c% reduction vary and are *h*^2^ = 12.2% (CI 95%, 5.2%-19.3%). However, we found that the 95% intervals of both estimates overlap, demonstrating that the estimates are relatively similar.

We further examined the stability of estimates between genders, splitting the cohort to males and females (36,098 and 38,773 patients respectively). Heritability estimates for absolute HbA1c% reduction in males were *h*^2^ = 15.9% (CI 95%, 1.2%-30.5%) and in women *h*^2^ = 20.9% (CI 95%, 7.5%-34.3%). Although the estimates are different, it is important to note the high overlap between the 95% CIs of the two groups, showing that the inter-heritability is comparable and is overall in the same relative values as the joint heritability measure.

We further investigated the heritability within the separated sexes group to the two additional phenotypes. We note that the Sci-LMM estimation never converged on the adjusted HbA1c% reduction for males, thus never yielding a concrete estimation. This is an artifact of the heuristic characteristic of the algorithm and does not indicate a lack of heritability for said measure.

We estimated heritability of responses to additional subgroups of the cohort, in order to search for affecting factors. We split our cohort by age (binning them by decades), and by absolute HbA1c% response values, and found no significant results (Supplementary Table 3). In addition we split the cohort by ethnicity, however, the resulting subgroups were all too small to provide estimates.

## Discussion

In this work, we estimated the heritability of response to metformin treatment in diabetic patients. Our cohort consists of 721,123 diabetic patients, 74,871 of whom begin metformin treatment while already recorded in the EHRs. In combination with pedigree information, we constructed a kinship matrix yielding genetic similarities between all patients. From it we estimated the heritable component of absolute reduction in HbA1c% following metformin in newly treated diabetic patients to be *h*^2^ = 10.5% (95% CI, 3.5%-17.5%). This value remained unchanged when adjusting the response for pretreatment personal covariates. In addition, we estimated for proportional HbA1c% reduction compared to baseline HbA1c% the heritability measure is *h*^2^ = 12.2% (95% CI, 5.2%- 19.3%).

Common approaches for estimating drug response heritability compute genetic similarities through genetic tests ^16,51-53^. Collecting genetic information leads to small cohorts and requires international collaborations. Our study obtained inherited similarities between individuals from pedigree information, these hold promise due to their tremendous size, and have been previously employed to estimate heritability measures of longevity, autism and others ^27,50^.

To the best of our knowledge, our study is the first to assess heritability of variation to drug response based on EHR data. We validated the proposed method by estimating heritability of adult height, finding the measure to be *h*^2^ = 80.7% (95% CI 80.3%-81.1%). This result agrees well with the widely accepted heritability of height of 80%^55^, thus strengthening our belief that our use of pedigrees in heritability estimates is robust to non-biological noise. Furthermore, it does not require costly genetic tests. We estimated the heritability of metformin responses to be in the range of 10%-20%, suggesting that while genetics likely contributes to variation in metformin glycaemic response for T2D patients, most of the variation is likely due to other environmental factors.

Estimated metformin responses heritability measures are within the parameters of previous genetic-based estimations, however with smaller confidence intervals ^16^. The increased statistical power is a direct result of our relatively large cohort size compared with previous works that commonly consist of up to several thousand patients ^16,52,53^.

Distinguishing between genetic and environmental effects is often difficult and not impossible. In our case, pedigree data encompasses more than just genetic information, as it provides some underlying information of environmental factors. Although this makes results more difficult to decipher, the accurate results of our positive and negative controls provide confidence in our method. We therefore believe that the included covariates capture the majority of the environmental variance and hence, prevent their effect on our *h*^2^ estimates. In spite of our efforts, we believe that it is still possible that some passed environmental information remains in our heritability estimates.

To ensure that we only account for the effect of metformin, we excluded from our cohort patients treated with other anti-diabetic drugs. However, we did not include covariates of other drugs that may interact with metformin and alter its effect. This is one of the limitations of working with EHR data compared to the much more controlled setting of randomized clinical trials. On the other hand, our analysis depicts real world scenarios and may thus provide more relevant estimates for the true underlying effect.

Our results show differences in the heritability of metformin responses by gender, with higher heritability measures among females. In spite of this, both measures have a shared 95% CI, thus reducing our confidence in difference between the sexes. We note that we also found significant differences in measurements at the time of treatment initialization, consistent with results from other countries ^56^. These differences in gender warrant further investigation, and call for a gender-specific disease management.

Predicting response to metformin for newly diagnosed patients holds great potential in diabetes treatment. Such predictors could help to faster divert non-responding patients to second line treatments with less deterioration. Several such predictors for second line treatments already exist, but most do not yield personalized recommendations ^57 58,59^. Our work suggests that predictors aiming to estimate metformin effects should probably include family medical history.

Overall, our results indicate that while genetics likely contribute to variation in metformin glycaemic response for T2D patients, environmental factors likely have a larger effect. Such findings are in line with prior evaluations of associations between single nucleotide polymorphisms and the reduction in HbA1c% after introduction to metformin ^11^. Our results emphasize the need for personalized treatment regimens of metformin. More generally, our work shows the utility of carrying out pharmacogenetic studies using EHRs, which may yield valuable insights without the burden and cost of genetic tests.

## Data Availability

The data that support the findings of this study originates from Clalit healthcare. Restrictions apply to the availability of these data and so are not publicly available. Due to restrictions, it can only be accessed through requests from the authors and/or the Clalit healthcare organization.

